# Dynamic liver function indexes monitoring and clinical characteristics in three types of COVID-19 patients

**DOI:** 10.1101/2020.05.13.20099614

**Authors:** Cheng Chen, Jie Jiang, Xiaoxiao Xu, Yiyang Hu, Yi Hu, Yu Zhao

## Abstract

**Background & Aims:** The abnormal liver function and even liver failure related death were reported in the COVID-19 patients, but less of studies focus on the dynamic liver function changes. We analysed the liver function indexes of COVID-19 patients to explore the characteristics of liver function changes in patients with different severity.

**Methods:** This study included 54 moderate, 50 severe, and 31 death nucleic acid-confirmed COVID-19 patients hospitalized at the central hospital of Wuhan, China. Epidemiological histories, clinical features, imaging materials, medications and especially major liver function laboratory tests were collected for analysis.

**Results:** The clinical symptoms did not present any significant difference in the patients at admission, but the older male patients had pronounced mortality risk. The normal ratio of ALT, TB, and DBIL of moderate patients was 96.3%, 94.44%, and 98.15% separately at the first test, but 59.26% of patients showed declined ALB levels. The normal ratio of all liver function indexes declined after admission, but most abnormalities were mild (1-2 times of upper limit unit) and went back normal before discharge. In severe patients, the normal ratio of ALB dropped down to 30.61% at admission along with the dramatic impaired normal ratio of bilirubin at the second test. The severe patients’ liver function dysfunction was worse than the moderate patients but without a significant difference. The dead patients showed a significantly higher level of DBIL, AST, GGT and CRP than other groups patients in the final test, along with the hypoalbuminemia. What is worse, 16.13% of non-survivors were diagnosed with liver failure. No medication was found to be related to ALT, AST, and GGT abnormality in our study.

**Conclusion:** In moderate and severe patients, liver dysfunction was mild. Patients widely presented lower level of ALB. The higher level of bilirubin, AST, and GGT was likely to indicate the worse outcome. Dynamic monitoring of liver function indexes could be considered and liver failure related death should be noticed and prevented in the early stage.

## Introduction

In December 2019, the first unknown pneumonia case which has been subsequently named coronavirus disease 2019 (COVID-19) was reported in Wuhan, China(1). To data, COVID-19 has become a pandemic disease warned by the World Health Organization (WHO) and triggered more than 3.8 million people in the world getting infected with this disease. The current global mortality of this disease nearly beat 6.94% (265,045 deaths among 3,820,156 confirmed cases, 14:30, 7^th^ May, Beijing time zone, Updating), even worse in some countries and states. It has become clear that COVID-19 patients are infected by the novel coronavirus SARS-CoV-2 which share a common ancestor with SARS-CoV(2). The following research suggested that angiotensin converting enzyme 2 (ACE2) is the main host cell receptor of SARS-CoV-2 and plays a crucial role in the entry of the virus into the cell to cause the final infection(2, 3). Since lung has a large number of type II alveolar cells which highly express ACE2, lung and respiratory system are the major targets of SARS-CoV-2, COVID-19 patients contain typical respiratory symptoms, such as dry cough and fever etc. However, dramatically changed liver function tests among the COVID-19 patients attracted more and more attention(4-10). One research from Shenzhen, China reported that 76.3% COVID-19 patients had abnormal liver test results and 90 in 419 patients had the liver injury during the hospitalization(4) Recently, a preliminary study showed that ACE2 receptor specifically expressed in cholangiocytes, indicating that virus might bind to ACE1-positive cholangiocytes and further led to cholangiocytes dysfunction(6). Although, there are increasing attention focusing on the liver damage in COVID-19 patient and two research pointed out the use of lopinavir/ritonavir can lead to the liver injury(4, 7), it is still unclear that how the liver function changes during the infection and whether these dysfunctions as the initial factor contributing to the progressing of this disease or as a result origin from the virus infection or/and the side effects because of the long term united medication. Furthermore, at present, it also keeps unknown whether the abnormal liver function or liver damage varies from different stages of patients.

Here, for monitoring the characteristics of liver function indexes and changes of COVID-19 patients. We collected clinical data, especially liver laboratory test and CRP test results from moderate, severe, and dead COVID-19 patients separately. And we dynamically analyzed the liver function indexes changes in these patients for providing reasonable and specific therapeutic strategies to the clinic.

## Results

### General information and clinical characteristics of COVID-19 Patients

All enrolled 135 COVID-19 patients came from Wuhan, China and hospitalized in the Central Hospital of Wuhan. The patients’ age distribution is from 23 years old to 99 years old. The patients’ age (*P*=0.007) and hospital stays (*P*<0.001) presented a significant difference. As the previous research showed, older, male patients are the most susceptible to SARS-CoV-2 infection(11), the ration of male patients went up from moderate to death group and presented the significant difference (*P* =0.001) in our study. The common symptoms among these patients were fever (95, 70.37%), dry cough (86, 63.7%), and fatigue (72, 53.33%) (**Table1**), several patients showed some digested symptoms and cardiovascular symptoms. But, all symptoms did not show any significant difference among the three groups. 23 dead patients (74.19%) once developed into acute respiratory distress syndrome (ARDS) during the hospitalization. Of the cause of death, 5 patients were diagnosed with liver failure (*P*<0.001).

**Table1.** General information and clinical characteristics of patients

| Characteristics | Moderate<br>N=54 | Severe<br>N=50 | Death<br>N=31 | P-value |
| --- | --- | --- | --- | --- |
| Male (%) | 24(44.44) | 27(54) | 27(87.1) | **0.001** |
| Age (years) | 58(23-91) | 63(28-83) | 66(34-99) | **0.007** |
| Smoking (%) | 4(7.41) | 5(10) | 4(12.9) | 0.654 |
| Drinking (%) | 2(3.7) | 6(12) | 3(9.68) | 0.284 |
| Hospitalization period<br>(days) | 24.5(3-87) | 36(6-78) | 15(1-67) | **<0.001** |
| Systemic symptoms (%) |  |  |  |  |
| Fever | 37(68.5) | 39(78) | 19(61.3) | 0.258 |
| Fatigue | 27(50) | 29(58) | 16(51.61) | 0.699 |
| Muscle pain | 13(24.07) | 17(34) | 4(12.9) | 0.101 |
| Respiratory symptoms (%) |  |  |  |  |
| Difficulty breathing | 4(7.41) | 2(4) | 5(16.13) | 0.167 |
| Cough | 37(68.52) | 33(66) | 16(51.61) | 0.27 |
| Sore throat | 2(3.7) | 5(10) | 2(6.45) | 0.406 |
| Wheezing | 11(20.37) | 17(34) | 13(41.94) | 0.09 |
| Gastrointestinal symptoms (%) |  |  |  |  |
| Abdominal pain | 1(1.85) | 5(10) | 0(0) | 0.108 |
| Diarrhea | 9(16.67) | 9(18) | 3(9.68) | 0.623 |
| Nausea | 6(11.11) | 3(6) | 2(6.45) | 0.666 |
| Vomiting | 2(3.7) | 2(4) | 0(0) | 0.677 |
| Circulatory symptoms (%) |  |  |  |  |
| Palpitations | 0(0) | 2(4) | 1(3.23) | 0.337 |
| Chest tightness | 6(11.11) | 5(10) | 8(25.81) | 0.135 |
| ARDS | 0(0) | 25(50) | 23(74.19) | **<0.001** |
| Shock | 1(1.85) | 2(4) | 10(32.26) | **<0.001** |
| Liver failure | 0(0) | 0(0) | 5(16.13) | **<0.001** |
| Antiviral Medication(%) |  |  |  |  |
| a-interferon | 8(14.81) | 9(18) | 2(6.45) | 0.375 |
| Lopinavir/ritonavir | 5(9.26) | 5(10) | 4(12.9) | 0.876 |
| Ribavirin | 46(85.19) | 45(90) | 25(80.65) | 0.484 |
| Arbidol | 41(75.93) | 30(60) | 8(25.81) | **<0.001** |
| Oseltamivir | 17(31.48) | 14(28) | 16(51.61) | 0.076 |
| Other therapeutic drugs(%) |  |  |  |  |
| Glucocorticoids | 28(51.85) | 39(78) | 26(83.87) | **0.002** |
| Antibiotics | 47(87.04) | 48(96) | 30(96.77) | 0.223 |
| Chinese medicine treatment | 50(92.59) | 44(88) | 15(48.39) | **<0.001** |
| Immunoglobulin | 18(33.33) | 29(58) | 17(54.84) | **0.027** |
| Liver protection | 11(20.37) | 12(24) | 8(25.81) | 0.828 |
| Breathing support(%) |  |  |  |  |
| Oxygen therapy (general) | 49(90.74) | 50(100) | 31(100) | **0.025** |
| High flow oxygen | 0(0) | 12(24) | 18(58.06) | **<0.001** |
| Non-invasive ventilator support | 0(0) | 6(12) | 11(35.48) | **<0.001** |
| Tracheal intubation | 0(0) | 3(6) | 6(19.35) | **0.001** |
| Prone position and lung expansion | 0(0) | 3(6) | 5(16.13) | **0.006** |
| ECMO | 0(0) | 3(6) | 4(12.9) | **0.017** |

NS=no statistics were calculated.

### Dynamic liver functional tests monitoring and analysis in moderate COVID-19 patients

Generally, the moderate patient showed an elevated systemic inflammation level but not typically in the liver at the initial stage of SARS-CoV-2 infection. Only slight liver damage or even mild abnormalities presented in several patients during the hospitalization. According to the first test results **(Table2)**, in the early stage of infection, the normal ratio of ALT, TB, DBIL, and IBIL was 96.3%, 94.44%, 98.15% and 90.74% separately, staying at a high level. 59.26% of patients showed slightly declined ALB levels at the beginning, indicating the mild dysfunction of hepatic protein synthesis. During the hospitalization, even the normal ratio of all liver function indexes declined, especially DBIL (*P*=0.0095), ALT (*P*=0.038), AST (*P*=0.044), and ALB (*P*=0.021) level significantly decreased, most patients just had a mild abnormality of these indexes (1-2 ULN raised in DBIL, ALT and AST; 0.75-0.5 LLN declined in ALB). However, it should be noticed that 27 patients’ first CRP test results were above 3 times the upper limit unit (ULN) (*P*<0.001) at admission **(Table2)**. The median of the first CRP test was 3.14 mg/dL, which was significantly higher (*P*<0.001) than the second and third test value. This dramatically increased CRP level indicated a higher systemic inflammatory response at the initial stage in moderate patients, but not especially happened in hepatocytes, considered with high level (96.3%) of normal ALT value. As hospital stays prolonging, the CRP level was getting a significant decline. The median of CRP decreased to 0.63 mg/dl, nearly 5 times lower than the first result and further went back to 0.155 mg/dl at the final test, which highly pointed out the recovery from the inflammatory disorders. Even these waxing and waning test results in the middle of hospitalization indicated slight inflammation and hepatic synthesis of protein dysfunction in COVID-19 patients, most abnormal function test went back to the normal at the final test.

**Table2.**
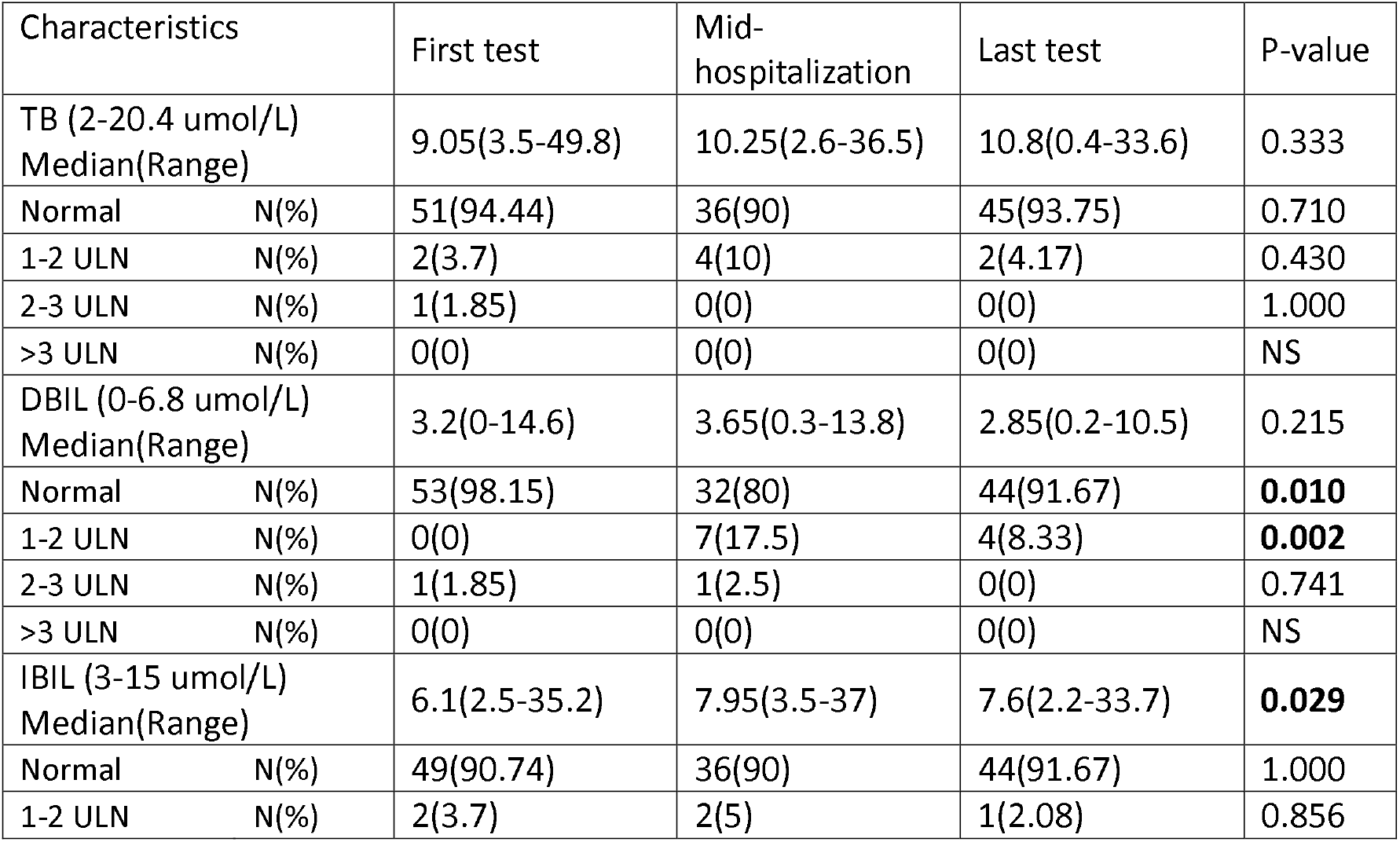
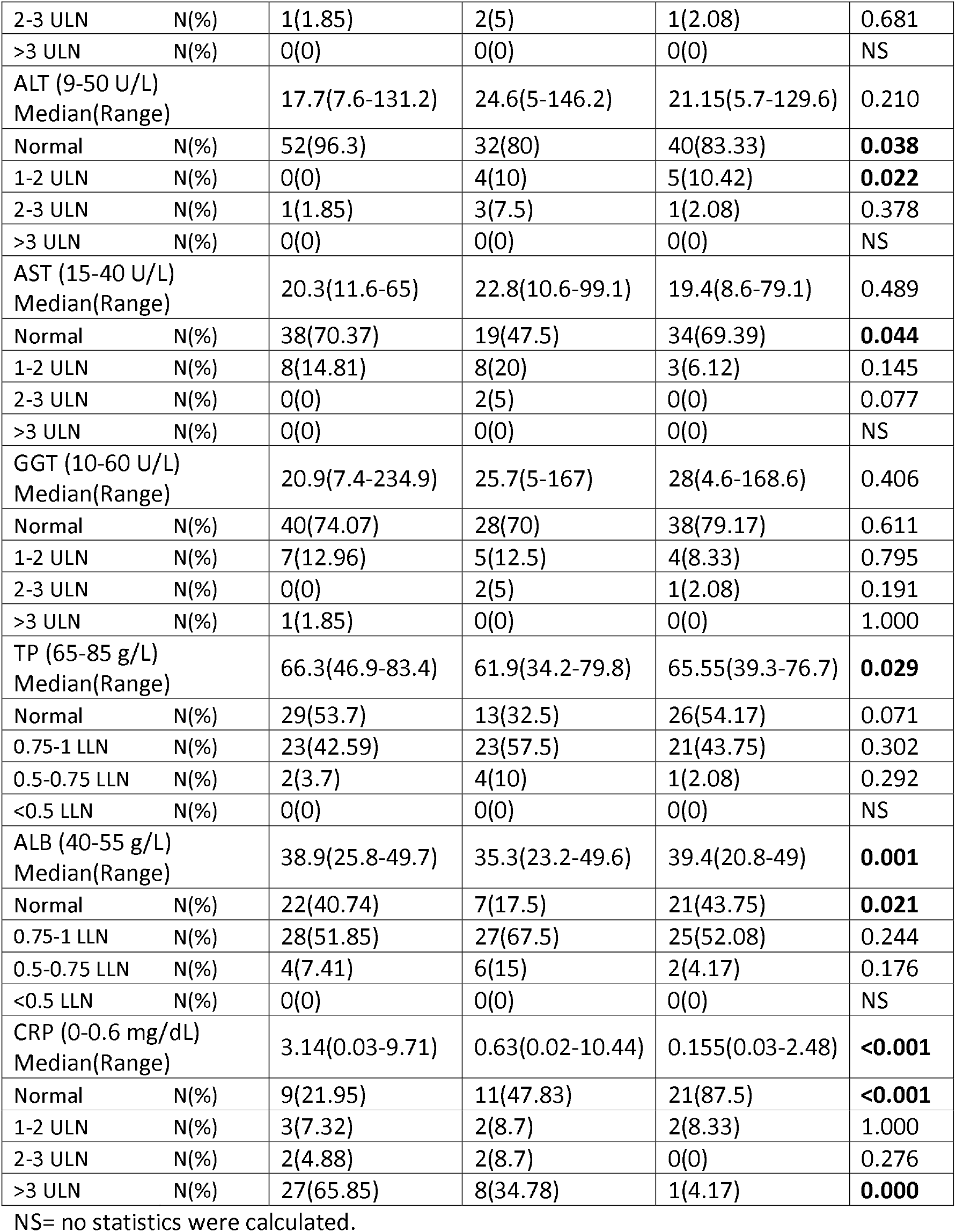
Dynamic liver functional tests analysis in moderate COVID-19 patients

| Characteristics | First test | Mid-hospitalization | Last test | P-value |
| --- | --- | --- | --- | --- |
| TB (2-20.4 umol/L)<br>Median(Range) | 9.05(3.5-49.8) | 10.25(2.6-36.5) | 10.8(0.4-33.6) | 0.333 |
| Normal N(%) | 51(94.44) | 36(90) | 45(93.75) | 0.710 |
| 1-2 ULN N(%) | 2(3.7) | 4(10) | 2(4.17) | 0.430 |
| 2-3 ULN N(%) | 1(1.85) | 0(0) | 0(0) | 1.000 |
| >3 ULN N(%) | 0(0) | 0(0) | 0(0) | NS |
| DBIL (0-6.8 umol/L)<br>Median(Range) | 3.2(0-14.6) | 3.65(0.3-13.8) | 2.85(0.2-10.5) | 0.215 |
| Normal N(%) | 53(98.15) | 32(80) | 44(91.67) | **0.010** |
| 1-2 ULN N(%) | 0(0) | 7(17.5) | 4(8.33) | **0.002** |
| 2-3 ULN N(%) | 1(1.85) | 1(2.5) | 0(0) | 0.741 |
| >3 ULN N(%) | 0(0) | 0(0) | 0(0) | NS |
| IBIL (3-15 umol/L)<br>Median(Range) | 6.1(2.5-35.2) | 7.95(3.5-37) | 7.6(2.2-33.7) | **0.029** |
| Normal N(%) | 49(90.74) | 36(90) | 44(91.67) | 1.000 |
| 1-2 ULN N(%) | 2(3.7) | 2(5) | 1(2.08) | 0.856 |

|  |  |  |  |  |  |
| --- | --- | --- | --- | --- | --- |
| 2-3 ULN | N(%) | 1(1.85) | 2(5) | 1(2.08) | 0.681 |
| >3 ULN | N(%) | 0(0) | 0(0) | 0(0) | NS |
| ALT (9-50 U/L) |  |  |  |  |  |
| Median(Range) |  | 17.7(7.6-131.2) | 24.6(5-146.2) | 21.15(5.7-129.6) | 0.210 |
| Normal | N(%) | 52(96.3) | 32(80) | 40(83.33) | **0.038** |
| 1-2 ULN | N(%) | 0(0) | 4(10) | 5(10.42) | **0.022** |
| 2-3 ULN | N(%) | 1(1.85) | 3(7.5) | 1(2.08) | 0.378 |
| >3 ULN | N(%) | 0(0) | 0(0) | 0(0) | NS |
| AST (15-40 U/L) |  |  |  |  |  |
| Median(Range) |  | 20.3(11.6-65) | 22.8(10.6-99.1) | 19.4(8.6-79.1) | 0.489 |
| Normal | N(%) | 38(70.37) | 19(47.5) | 34(69.39) | **0.044** |
| 1-2 ULN | N(%) | 8(14.81) | 8(20) | 3(6.12) | 0.145 |
| 2-3 ULN | N(%) | 0(0) | 2(5) | 0(0) | 0.077 |
| >3 ULN | N(%) | 0(0) | 0(0) | 0(0) | NS |
| GGT (10-60 U/L) |  |  |  |  |  |
| Median(Range) |  | 20.9(7.4-234.9) | 25.7(5-167) | 28(4.6-168.6) | 0.406 |
| Normal | N(%) | 40(74.07) | 28(70) | 38(79.17) | 0.611 |
| 1-2 ULN | N(%) | 7(12.96) | 5(12.5) | 4(8.33) | 0.795 |
| 2-3 ULN | N(%) | 0(0) | 2(5) | 1(2.08) | 0.191 |
| >3 ULN | N(%) | 1(1.85) | 0(0) | 0(0) | 1.000 |
| TP (65-85 g/L) |  |  |  |  |  |
| Median(Range) |  | 66.3(46.9-83.4) | 61.9(34.2-79.8) | 65.55(39.3-76.7) | **0.029** |
| Normal | N(%) | 29(53.7) | 13(32.5) | 26(54.17) | 0.071 |
| 0.75-1 LLN | N(%) | 23(42.59) | 23(57.5) | 21(43.75) | 0.302 |
| 0.5-0.75 LLN | N(%) | 2(3.7) | 4(10) | 1(2.08) | 0.292 |
| <0.5 LLN | N(%) | 0(0) | 0(0) | 0(0) | NS |
| ALB (40-55 g/L) |  |  |  |  |  |
| Median(Range) |  | 38.9(25.8-49.7) | 35.3(23.2-49.6) | 39.4(20.8-49) | **0.001** |
| Normal | N(%) | 22(40.74) | 7(17.5) | 21(43.75) | **0.021** |
| 0.75-1 LLN | N(%) | 28(51.85) | 27(67.5) | 25(52.08) | 0.244 |
| 0.5-0.75 LLN | N(%) | 4(7.41) | 6(15) | 2(4.17) | 0.176 |
| <0.5 LLN | N(%) | 0(0) | 0(0) | 0(0) | NS |
| CRP (0-0.6 mg/dL) |  |  |  |  |  |
| Median(Range) |  | 3.14(0.03-9.71) | 0.63(0.02-10.44) | 0.155(0.03-2.48) | **<0.001** |
| Normal | N(%) | 9(21.95) | 11(47.83) | 21(87.5) | **<0.001** |
| 1-2 ULN | N(%) | 3(7.32) | 2(8.7) | 2(8.33) | 1.000 |
| 2-3 ULN | N(%) | 2(4.88) | 2(8.7) | 0(0) | 0.276 |
| >3 ULN | N(%) | 27(65.85) | 8(34.78) | 1(4.17) | **0.000** |

NS= no statistics were calculated.

### Dynamic liver functional tests monitoring and analysis in severe COVID-19 patients

The normal ratio of AST level (59.18%), TP level (67.35%), especially the ALB (30.61%) and CRP (17.39%) levels of severe patients were dramatic low at the first test **(Table3)**. In the middle of hospitalization, there were pronounced increased TB, DBIL, IBIL, ALT (from 21.1 U/L to 37.45 U/L, *P*=0.028) levels, and decreased ALB level. The test results indicated hepatocytes damage or hepatitis happened in some severe COVID-19 patients during the hospitalization. What is more, the liver damage among the severe patients tended to be symbolized with significant elevated TB, DBIL and IBIL level in the early period of the hospitalization. However, the median of AST attenuated significantly (*P*=0.011) after admission. And the median of CRP has been significantly decreased (*P*=0.003) during the hospitalization.

**Table3.**
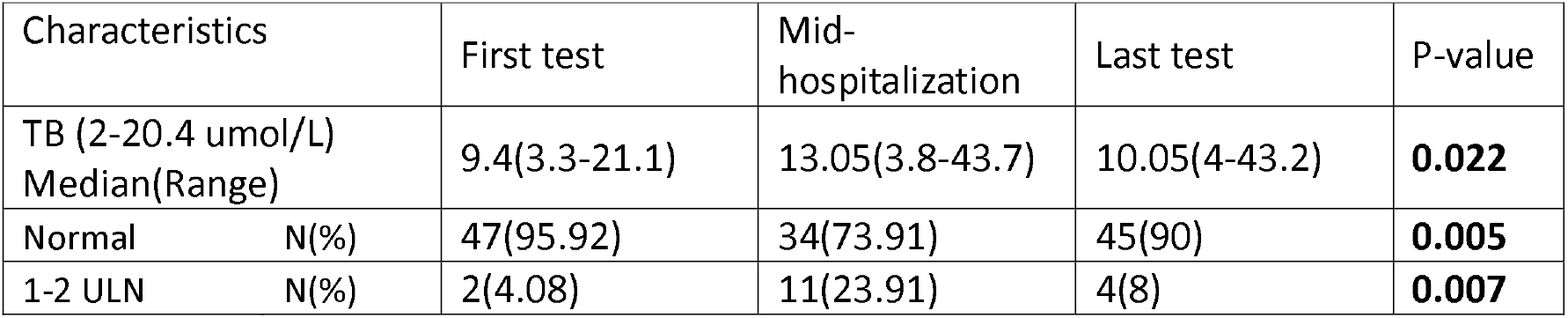
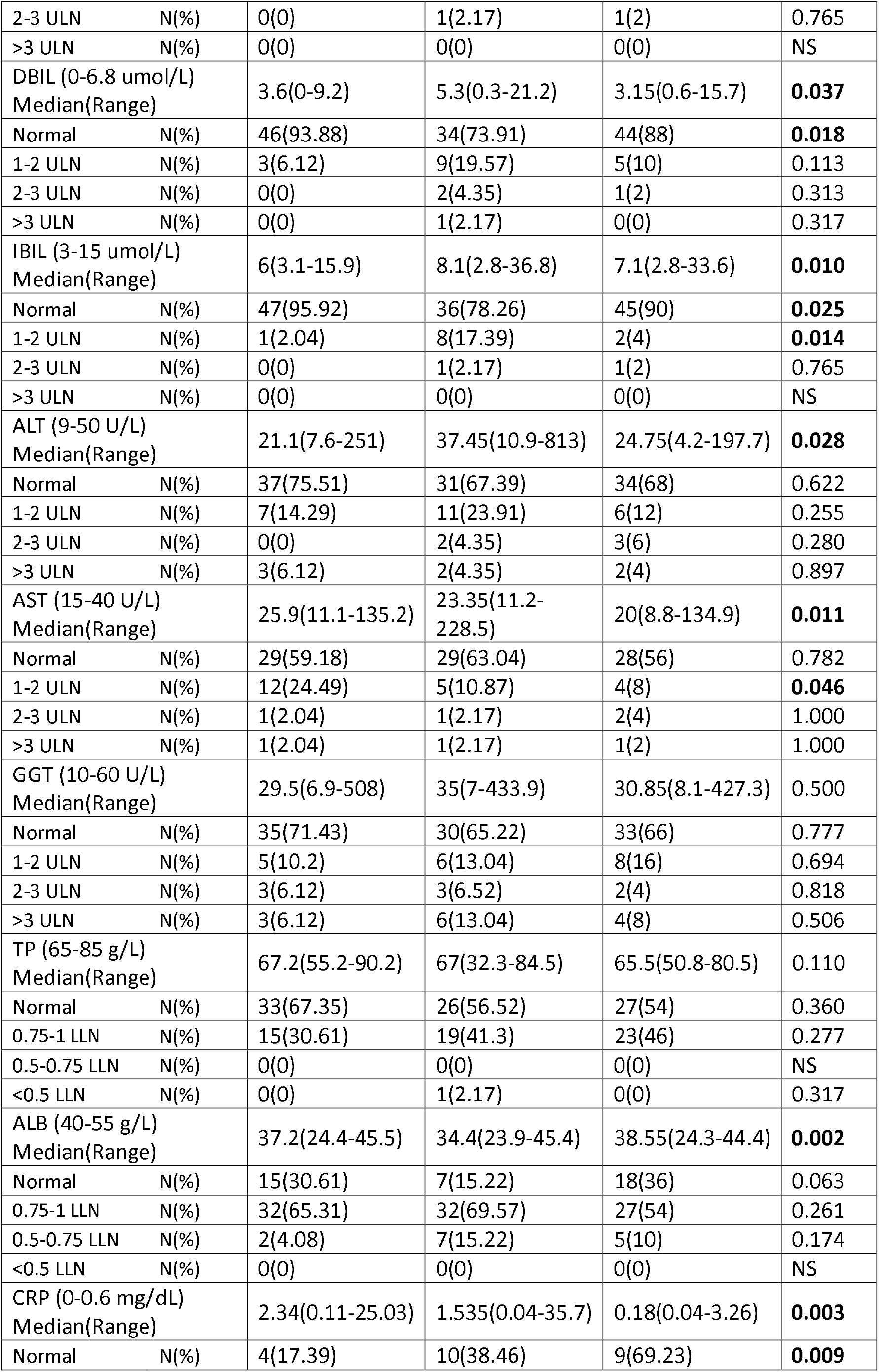
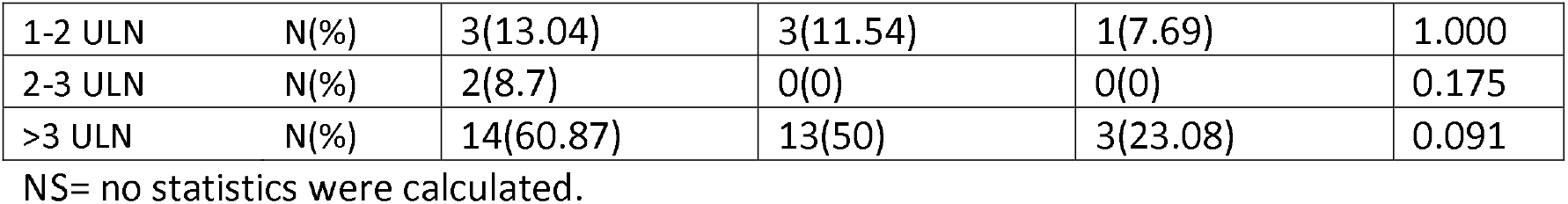
Dynamic liver functional tests analysis in severe COVID-19 patients

### Dynamic liver functional tests monitoring and analysis in dead COVID-19 patients

The normal ratio of DBIL, ALT, AST and GGT had a persistently dropped tend during the hospitalization **(Table 4)**. GGT was considered as a sensitive laboratory index of cholangiocytes dysfunction. Taking consideration of the ALT, AST, and ALB levels were persistent sinking in during the hospitalization, the worse liver function test results showed the increased liver inflammation and liver injury happened in some COVID-19 patients. Similarly, ALB value kept the lower level. Seventeen (54.84%) and 12 (38.71%) patients at the third test had 0.75 times (*P*=0.015) and 0.5 times (*P*=0.001) below than the LLN separately, which highly presented a much worse hepatic synthesis of protein dysfunction. The patients died during the hospitalization were accompanied by a persistent abnormal and up-going CRP level, even no patient presents a normal CRP level at admission. The last median of CRP was 5.89 mg/dl **(Table 4)**, the highest value at all the observation time point. indicating the progressive inflammatory response before death. Considering 5 liver failure cases were reported, there was a higher incidence of acute liver injury and rapidly deteriorated liver functions including impaired hepatic synthesis of protein, hepatic inflammation and injury in COVID-19 patients.

**Table4.** Dynamic liver functional tests analysis in dead group of COVID-19 patients

| Characteristics |  | First test | Mid-hospitalization | Last test | P-value |
| --- | --- | --- | --- | --- | --- |
| TB (2-20.4 umol/L)<br>Median(Range) |  | 9.6(2.6-30.3) | 15.9(3.4-44.7) | 13.7(1.6-281.2) | 0.058 |
| Normal | N(%) | 24(82.76) | 12(66.67) | 22(70.97) | 0.405 |
| 1-2 ULN | N(%) | 5(17.24) | 5(27.78) | 7(22.58) | 0.687 |
| 2-3 ULN | N(%) | 0(0) | 1(5.56) | 0(0) | 0.231 |
| >3 ULN | N(%) | 0(0) | 0(0) | 1(3.23) | 1.000 |
| DBIL (0-6.8 umol/L)<br>Median(Range) |  | 4(0-12) | 6.25(2-15.7) | 5.8(0-201.2) | **0.046** |
| Normal | N(%) | 23(79.31) | 12(66.67) | 18(58.06) | 0.210 |
| 1-2 ULN | N(%) | 6(20.69) | 4(22.22) | 10(32.26) | 0.606 |
| 2-3 ULN | N(%) | 0(0) | 2(11.11) | 2(6.45) | 0.215 |
| >3 ULN | N(%) | 0(0) | 0(0) | 1(3.23) | 1.000 |
| IBIL (3-15 umol/L)<br>Median(Range) |  | 7.9(1.1-19.5) | 8.9(4.4-32.2) | 7.7(0.2-80) | 0.177 |
| Normal | N(%) | 22(75.86) | 14(77.78) | 23(74.19) | 1.000 |
| 1-2 ULN | N(%) | 2(6.9) | 2(11.11) | 4(12.9) | 0.805 |
| 2-3 ULN | N(%) | 0(0) | 1(5.56) | 0(0) | 0.231 |
| >3 ULN | N(%) | 1(3.45) | 0(0) | 1(3.23) | 1.000 |
| ALT (9-50 U/L)<br>Median(Range) |  | 21.1(6.2-62.2) | 20.3(7-92.7) | 27(6.2-645.9) | 0.100 |
| Normal | N(%) | 25(86.21) | 14(77.78) | 23(74.19) | 0.522 |
| 1-2 ULN | N(%) | 2(6.9) | 2(11.11) | 3(9.68) | 0.888 |
| 2-3 ULN | N(%) | 0(0) | 0(0) | 0(0) | NS |
| >3 ULN | N(%) | 0(0) | 0(0) | 3(9.68) | 0.176 |
| AST (15-40 U/L)<br>Median(Range) |  | 31(9-312) | 32.1(14-136.7) | 33(9-640.6) | 0.262 |
| Normal | N(%) | 18(62.07) | 12(66.67) | 16(51.61) | 0.535 |
| 1-2 ULN | N(%) | 4(13.79) | 3(16.67) | 7(22.58) | 0.701 |
| 2-3 ULN | N(%) | 1(3.45) | 0(0) | 3(9.68) | 0.442 |
| >3 ULN | N(%) | 1(3.45) | 1(5.56) | 3(9.68) | 0.839 |
| GGT (10-60 U/L)<br>Median(Range) |  | 34.8(11.4-133.2) | 33.15(10.9-172.9) | 44(9.7-143) | 0.288 |
| Normal | N(%) | 24(82.76) | 14(77.78) | 20(64.52) | 0.274 |
| 1-2 ULN | N(%) | 3(10.34) | 2(11.11) | 6(19.35) | 0.648 |
| 2-3 ULN | N(%) | 2(6.9) | 2(11.11) | 4(12.9) | 0.805 |
| >3 ULN | N(%) | 0(0) | 0(0) | 0(0) | NS |
| TP (65-85 g/L)<br>Median(Range) |  | 67(55-82) | 66.25(54-91.7) | 67(47.3-104.9) | 0.879 |
| Normal | N(%) | 19(65.52) | 8(44.44) | 17(54.84) | 0.357 |
| 0.75-1 LLN | N(%) | 10(34.48) | 8(44.44) | 11(35.48) | 0.765 |
| 0.5-0.75 LLN | N(%) | 0(0) | 0(0) | 1(3.23) | 1.000 |
| <0.5 LLN | N(%) | 0(0) | 0(0) | 0(0) | NS |
| ALB (40-55 g/L) |  |  |  |  |  |
| Median(Range) |  | 36.1(28.1-45.1) | 30.2(24.8-46.2) | 32.6(22.6-45.1) | 0.000 |
| Normal | N(%) | 4(13.79) | 2(11.11) | 2(6.45) | 0.639 |
| 0.75-1 LLN | N(%) | 24(82.76) | 8(44.44) | 17(54.84) | **0.015** |
| 0.5-0.75 LLN | N(%) | 1(3.45) | 7(38.89) | 12(38.71) | **0.001** |
| <0.5 LLN | N(%) | 0(0) | 1(5.56) | 0(0) | 0.231 |
| CRP (0-0.6 mg/dL) |  |  |  |  |  |
| Median(Range) |  | 5.19(0.96-21.92) | 3.545(0.78-26.98) | 5.89(2.06-25.38) | 0.443 |
| Normal | N(%) | 0(0) | 0(0) | 0(0) | NS |
| 1-2 ULN | N(%) | 1(4.17) | 1(10) | 0(0) | 0.466 |
| 2-3 ULN | N(%) | 1(4.17) | 2(20) | 0(0) | 0.128 |
| >3 ULN | N(%) | 22(91.67) | 7(70) | 18(100) | 0.038 |

NS=no statistics were calculated.

### The difference of liver function tests and characteristics between three types of COVID-19 patients

For identifying the differences among these three types of patients, we further compared the liver function indexes among these patients at each observation time point. At the first test, the CRP level in non-survivors was dramatically higher than the level in other two groups, but the value distribution of TB, DBIL, ALT, TP, and ALB did not show any pronounced difference among three types of patients. The AST (*P*=0.026) and GGT (*P*=0.013) **(Sup.1)** were gradually increased from moderate to dead group patients and the expression of moderate patients was significantly lower **(Fig.1)**. At the second test, there was no significant difference in the distribution of liver function indexes and CRP value between moderate and severe patients. But the median of CRP (3.545 mg/dL) in dead patients was significantly higher, as well as TB (15.9 umol/L) and DBIL (6.25 umol/L) value. Meanwhile, there was a dramatic low median of ALB (30.2 g/L) in dead patients. Besides, ALT went down to the bottom and showed a significantly low level (20.3 U/L) in dead patients group. Even GGT and AST did not show the pronounced difference among three groups patients, but they significantly went up in the dead patients group in the final test (44U/L and 33U/L, separately).

**Fig.1.**
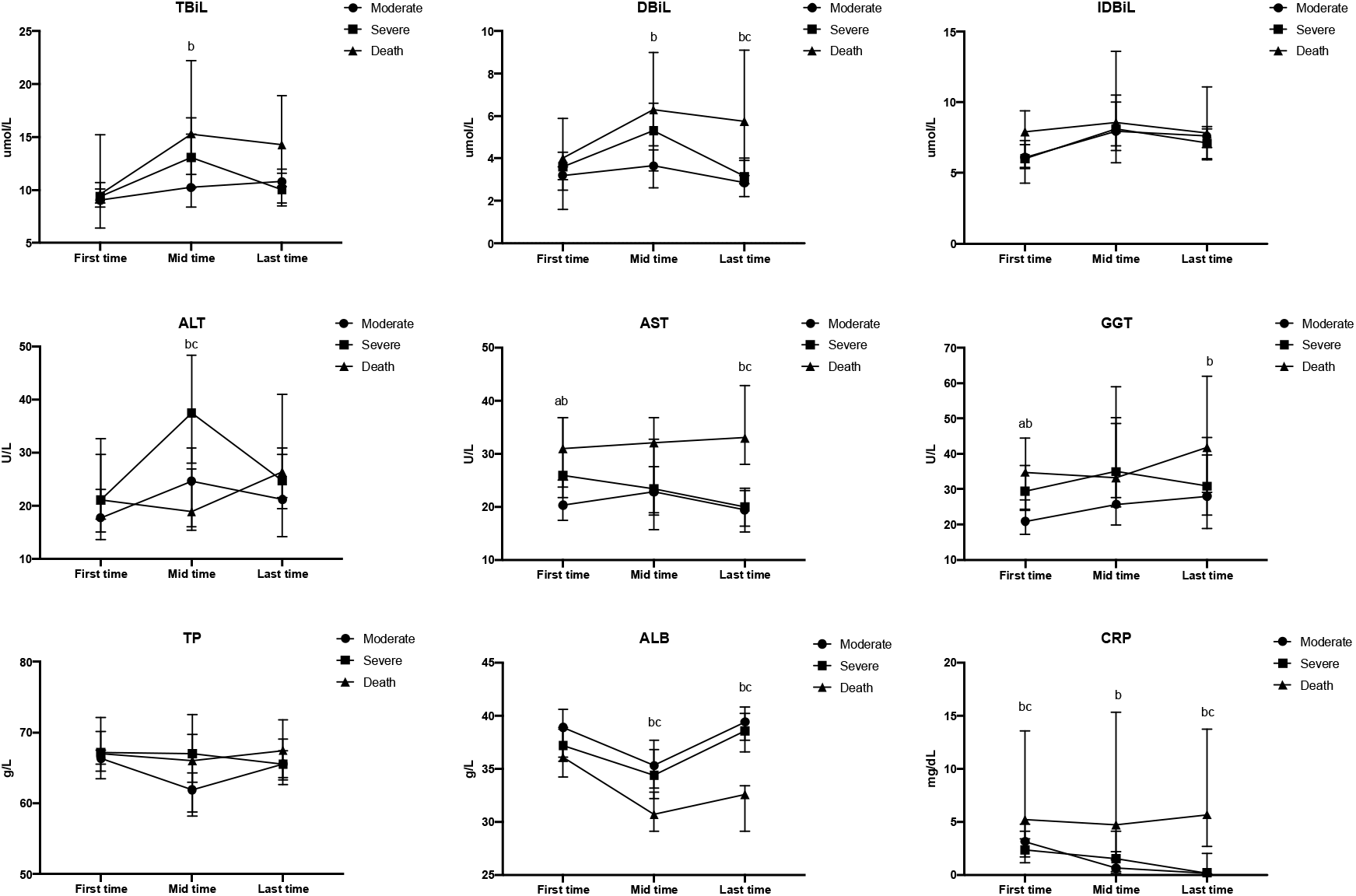
The changes of liver function indexes and CRP in COVID-19 patients. The variables were expressed as median and 95% CI. First time: the first test; Mid time: in the middle test of hospitalization; Last time: the last test. a: Sever group vs Moderate group; b: Death group vs Moderate group; c: Sever group vs Death group. abc *P*<0.05.

Taken together, from the admission to discharge, the liver function changes were not significantly different between moderate and severe patients, but the persistently high level of DBIL, AST and continuing impaired ALB synthesis in dead patients pointed out that these liver function indexes changes were more sensitive and typical to reflect the worse outcome of the COVID-19 patients.

### The medication effect on liver function analysis in COVID-19 patients during the hospitalization

During the hospitalization, the most used top three antiviral drugs were ribavirin, arbidol, and oseltamivir **(Table1)**. There was a significant statistical difference (*P*<0.001) in arbidol usage between dead patients and the other two group patients. The usage of antibiotics got increased with the progress of the disease, from 87.04% in moderate patients to 96% and 96.77% in severe and dead patients. Specially, traditional Chinese herbs were widely used among moderate (92.59%) and severe (88%) patients. Most COVID-19 patients got the antiviral treatment after admission, for analyzing the effects of medications on liver function in these patients, we chose 104 patients who showed the normal liver ALT and AST, 109 patients who had normal liver GGT level at admission but presented the liver abnormalities during the hospitalization for data analysis **(Table5)**. We adjusted potential confounding by age, gender, ARDS, liver failure, shock and liver comorbidities according to table1, Interestingly, glucocorticoids (OR=0.073, 95% CI 0.012-0.438, P=0.004) and Chinese herbs (OR= 0.151, 95% CI 0.027 - 0.836, P=0.03) presented pronounced protective effects on ALT or AST abnormality. And glucocorticoids had significant protective effect on GGT change (OR= 0.046, 95% CI 0.005-0.444, P=0.008). All antiviral drugs including lopinavir/ritonavir, ribavirin, arbidol, and oseltamivir did not show any significant effect on ALT, AST, or GGT abnormalities.

**Table 5.** The medication of COVID-19 patients after admission

|  | ALT or AST |  |  | GGT |  |  |
| --- | --- | --- | --- | --- | --- | --- |
|  | Adjusted OR | (95%CI) | P-value | Adjusted OR | (95%CI) | P-value |
| a-interferon | 5.165 | (0.803, 33.242) | 0.084 | 1.962 | (0.372, 10.353) | 0.427 |
| Lopinavir/ritonavir | 1.943 | (0.21, 17.936) | 0.558 | 0.331 | (0.034, 3.23) | 0.341 |
| Ribavirin | 0.085 | (0.007, 1.112) | 0.06 | 0.438 | (0.039, 4.887) | 0.502 |
| Arbidol | 1.303 | (0.369, 4.606) | 0.681 | 0.75 | (0.21, 2.671) | 0.657 |
| Oseltamivir | 0.687 | (0.197, 2.402) | 0.557 | 1.277 | (0.378, 4.314) | 0.694 |
| Glucocorticoids | 0.073 | (0.012, 0.438) | 0.004 | 0.046 | (0.005, 0.444) | 0.008 |
| Antibiotics | 13.355 | (0.69, 258.538) | 0.086 | 1.135 | (0.133, 9.719) | 0.908 |
| Chinese herbs | 0.151 | (0.027, 0.836) | 0.03 | 1.786 | (0.399, 8.003) | 0.449 |

## Discussion

In our present study, we separately and dynamically analyzed the liver function tests at three-time points of three types of COVID-19 patients hospitalized in the Central Hospital of Wuhan, China. To the best of our knowledge, this study is the first one to dynamically analyze the liver function changes and characteristics in COVID-19 patients ranging from the moderate stage to death.

It has become clear that moderate patients, severe patients, and the patients died during the hospitalization demonstrate separate but typical characteristics. Generally, except widely impaired ALB and TP levels in COVID-19 patients at admission, more than 70% patient presented normal liver injury indexes at the initial stage, which consistent with a previous study(7). And only a small number of moderate COVID-19 patients has a mild increase in liver enzymes (1-2 UNL) during the hospitalization. In severe patients, the median of GGT, especially ALT (from 21.1 U/L to 37.45U/L) presented an elevated level at the second test. Considering the higher TB and DBIL medians, the severe patients are highly present developing hepatocytes inflammation as well as liver injury in the early period of hospitalization. However, both moderate and severe patients, the abnormalities of the inflammatory index and liver enzymes showed a very clear and undoubted tendency to go back normal in the end. Unexpectedly, there is a more normal ratio of ALT, GGT, and AST level in dead group patients than the severe group at the beginning, but the CRP (*P*=0.030) and ALB (*P*=0.041) normal ratio are dramatically lower than the other two groups, which demonstrated a higher level of the inflammatory response and impaired hepatic protein synthesis but not a wide spectrum of liver dysfunction happened in the dead patients at the initial stage. However, these high levels of the inflammatory response and impaired hepatic protein synthesis keep until patients’ death. The median of DBIL, AST, and GGT kept pronounced high level after admission in the non-survivors, which are more likely to be related to the worse outcome of the COVID-19 patients. In our study, 5 liver failure diagnosis reported in these dead patient, 3 of them presented total normal liver function tests except abnormal ALB or/and TP level at admission but showed extremely elevated ASL, ALT or/and GGT value before death. These severe liver function derangements in these three patients indicate different liver damage situation. After very carefully data tracking and comparison, their liver function derangements happened in a very short period and they ended up with an acute liver injury or liver failure. However, it should be pointed out that, except the lower ALB and TP levels, 10 patients (32.25%) in the dead group nearly did not show any abnormality in liver function test before death and increasing evidence pointed out that liver failure is not a main concern and liver is not the target of significant inflammatory damage in COVID-19 patients(12).

Undeniably, liver dysfunction is happening in COVID-19 patients. A postmortem biopsy of the patient with COVID-19 performed in China showed that moderate microvascular steatosis and mild lobular and portal activity presented in the liver tissue(13). Similarly, another postmortem wedge liver biopsies research from Italy pointed out that minimal inflammation features showed in the liver tissue of COVID-19 patients without significant liver disease complain or signs of liver failure before death(12). But as the postmortem biopsy showed, most derangement of liver damage is clearly mild(12-14). Based on our dynamically analyzed results, only a few patients showed pronounced (>2 ULN or <0.75LLN) changes in liver laboratory tests from the admission to discharge. Even one research demonstrated that cholangiocytes express ACE2 suggesting a potential mechanism of direct damage to the hepatobiliary system in SARS-CoV-2 infection(6). Like Qingxiao et al. (4) proved that unpronounced ALP level in most patients indicated the safety of cholangiocytes since ALP is more sensitive than GGT to reflect bile duct injury. Similarly, only a few of severe patients showed elevated ALP value in our study (data not shown).

There is emerging evidence to partly explain the liver injury in COVID-19 patients. In human small intestinal organoids (hSIOs), enterocytes were readily infected by SARS-CoV-2, which experimental released that intestine may present the viral target organ(15). This potential mechanism could be an explanation of the diarrhoea and gastrointestinal symptoms happened in COVID-19 patients(16). Considering the close pathophysiological connection between intestine and liver, add up with antibiotics widely be used in COVID-19 patient, the intestinal dysfunction might be happened and further aggravate the liver derangement. Meanwhile, vascular portal and sinusoidal thrombosis were observed in COVID-19 patients(12), which lead to focal parenchymal necrosis and liver cells accelerated apoptosis. And this identify was consistent with the presence of antiphospholipid antibodies in COVID-19 patients(17), indicating the multifocal thrombosis in critical patients. On the other hand, based on our data, the usage of the drugs during the hospitalization is not a real risk factor to the liver injury in COVID-19 patients. Consistent with two previous research(4, 7), ribavirin, arbidol, and oseltamivir do not lead to liver derangement in COVID-19 patients. Interestingly, Chinese herbs were proved to be safe to the liver function of COVID-19 patients(4), furthermore, the Chinese herbs even showed the significant protective effect to prevent ALT or AST abnormality in our study. But, we do not re-confirm the result that lopinavir/ritonavir increased the odds of liver injury like the previous said, that would be attributed to the poor usage ratio of lopinavir/ritonavir is these 135 patients, we cannot adequately analyze this risk factor.

Besides, the dramatically increased CRP level at the first test among the three types of patients indicated a severe inflammatory response happening in the early stage of virus infection. This universal elevated CRP level in these patients consistent with a previous study, which pointed out that increased CRP level was highly correlated with acute lung injury in COVID-19 patients (18). However, this inflammatory response is gradually revised in moderate and severe patients. The continuous rise in CRP level is highly correlated with the cytokine storm and a series of immune responses in dead patients (1, 19). Furthermore, the liver damage presented in some of the COVID-19 patients is highly due to a dysregulated inflammatory response because of the virus infection rather than to the presence of specific and severe underlying liver disease(14, 21, 22). On the contrary, ALB is negatively correlated with the severity of COVID-19 patients in our study. Although a mild decline of ALB level showed in moderate and severe patients at admission, the majority present a tendency to go up to the normal or stop going worse during the hospitalization. Some previous studies also showed that hypoalbuminemia was a potent, dose-dependent independent predictor of poor clinical outcomes(20) and ALB may be a negative predictor of the COVID-19 severity(18). Even one research believed that impaired albumin is more likely due to the COVID-19 patients’ consumption and inadequate protein intake caused by poor appetite(19). The declined ALB level widely presented nearly 60% of patients at admission in our study, which is highly attributed to the impaired synthesis of protein function in the liver.

To the limitations of this study, the number of patients in each group in the present study is limited, and all data were collected from a single hospital, a large scale study in multi-centre, even in multi-country could be designed in the future. On the other hand, we did not systemically consider the whole disease situation basing on other related laboratory tests, that would be harder for us to take consideration of the liver injury to the whole disease progression in the patients.

In conclusion, in our study, we separately and dynamically analyzed the liver function tests at three-time points of moderate, severe and dead COVID-19 patients. Liver dysfunction and damage truly happened in some COVID-19 patients and most liver derangement and damage happened and further developed after admission. Severe acute liver injury and failure presented in patients, which should be noticed and prevented in the early stage to help decreasing mortality. However, most hepatic abnormalities are mild and are more likely to be the subsequent consequences of systemic inflammatory dysfunction in COVID-19 patients, more attention should be paid to focus towards viral control and modulating systemic inflammation in COVDI-19 patients.

## Methods

### Patients

This is a single-centre, retrospective, cross sectional study, 135 COVID-19 patients who confirmed by SARS-CoV-2 nucleic acid in the Central Hospital of Wuhan, China, admitted from December 5, 2019, to January 31, 2020, were included. The patients’ exposure history, clinical symptoms, laboratory examinations and chest CT scans images also have been considered. The study was approved by the Institutional Review Boards of Hubei Public Health Clinical Center, the Central Hospital of Wuhan, China.

According to the coronavirus pneumonia diagnosis and treatment plan (trial version 7) developed by the National Health Committee of the People’s Republic of China, we classified the surviving patients into the moderate group or severe group, and death cases were classified as death group. The moderate (non-severe) type was defined as COVID-19 cases with symptoms and not severe pneumonia on radiological images. Severe pneumonia was defined of any of the followings: (1) respiratory distress, the respiratory rate per min≥30; (2) in the resting state, means oxygen saturation≤93%; (3) arterial blood oxygen partial pressure/oxygen concentration≤300mmHg (1mmHg = 0.133kPa); (4) progress of chest radiological manifestations> 50% within 24-48 hours. Finally, 54 patients were grouped into moderate patients, 50 patients were grouped into severe patients and rest (31 cases) belonged to the dead group.

### Data collection

The collected data were provided by the Central Hospital of Wuhan, including demographics data, clinical symptoms, chest CT scans, medication and treatment, and independent liver functional laboratory results during the hospitalization. We separately analyzed the liver function indexes at the first test, in mil-hospitalization (the second test), and the last test (the third test). Total bilirubin (TB), Direct bilirubin (DBIL), Indirect bilirubin (IBIL), Alanine transaminase (ALT), Aspartate transaminase (AST), Gamma-glutamyltransferase (GGT), Total protein (TP), Albumin (Alb) and C-reaction protein (CRP) were included. Since some moderate and dead patients had short hospitalization period, they only accepted two laboratory tests, we defined as the first and third test. A few of them only got one test, we defined as the first test. All data were acquired by physicians in the Central Hospital of Wuhan and further checked the accuracy by the other two researchers.

### Statistical analysis

Continuous variables were expressed as median (range, R) and compared with the Wilcoxon H test, and the all pairwise adjusted P-value was adopted for *post-hoc* multiple comparisons.; categorical variables were expressed as number (%) and compared by χ2 test or Fisher’s exact test if appropriate. Multiple logistic regression was used to explore the association between drugs and liver injury, selection of covariates was based on the clinical baseline comparison of different groups of COVID-19 Patients A two-sided α of less than 0.05 was considered statistically significant. Statistical analyses were done using SPSS version 24.0 (IBM SPSS, Armonk, NY).

## Data Availability

All data referred to in the manuscript is the availability

## Abbreviation

COVID-19: coronavirus disease 2019
SARS-CoV-2: severe acute respiratory syndrome coronavirus 2
ACE2: angiotensin converting enzyme 2
ARDS: acute respiratory distress syndrome
TB: total bilirubin
DBIL: direct bilirubin
IBIL: indirect bilirubin
ALT: alanine transaminase
AST: aspartate transaminase
GGT: gamma-glutamyltransferase
TP: total protein
ALB: albumin
CRP: C-reaction protein
ULN: upper limit unit
LLN: lower limit unit

## Author contributions

YZ, CC designed this study. JJ, XX, and YH collected and collated the data. YZ, YYH analysed the data. CC wrote the manuscript. YZ revised the manuscript. All authors critically participated in the discussion and commented on the manuscript.

## Acknowledgements

Thanks to all the clinical staff who is working extremely hard to fight this horrible disease in the world. We will win against the COVID-19. (CC is grateful to the financial support of the China Scholarship Council.)

